# COVID19 Real-World Data for the US and Lessons to Re-Open Business

**DOI:** 10.1101/2020.06.02.20120618

**Authors:** Pascal J. Goldschmidt-Clermont

## Abstract

In March of 2020, the COVID19 pandemic had expanded to the United States of America (US). Companies designated as “essential” for the US had to maintain productivity in spite of the growing threat created by the SARS-CoV-2 virus. With this report, we present the response of one such company, the Lennar Corporation, a major homebuilder in the US. Within days, Lennar had implemented a morning health check via its enterprise resource planning system, to identify associates (employees) who were sick, or not in their “usual state of health”. With this survey, Lennar was able to ensure that no one sick would show up to work, and instead, would self-quarantine at home. Furthermore, with thorough contact tracking, associates exposed to COVID19 patients (suspected or RT-PCR test-confirmed), were also asked to self-quarantine. This survey, in addition to other safety measures, such as an overhaul of the company with nearly 50% of the company working from home, prolific communication, and many more measures, Lennar was able to function safely for its associates and successfully as an enterprise. The data that we present here are “real world data” collected in the context of working throughout a dreadful pandemic, and the lessons learned could be helpful to other companies that are preparing to return to work.

## INTRODUCTION

SARS-CoV-2 became a human health threat during the second half of 2019 starting in the Wuhan city of China, and by the first quarter of 2020, led to a global pandemic (1). SARS-CoV-2 (CoV-2) can cause Acute Respiratory Distress Syndrome (ARDS), and seems to be more contagious than SARS-CoV-1 (2). Until recently, our information on the prevalence of the pandemic was limited to data collected on people who were tested and were positive for COVID-19 (1,551,095 total cases on May 21, 2020), and then amongst those positive individuals, patients who died (93,061 total death on May 20, 2020) (3,4).

While helpful in justifying the “stay at home order”, a potential limitation of such data, is that it was impacted markedly by guidelines (local and national) on who could be tested (5), and in the US, our real-world experience is that the majority of individuals who have presented with typical COVID-19 symptoms could not be tested because of limited supply of test kits. Indeed, Bendavid *et al*. have shown that by early April, seroprevalence of antibodies specific for CoV-2 for a cohort of volunteers recruited in Santa Clara County, California, was much greater than the number of confirmed cases reported for the same region (6). Furthermore, because the bias for testing appears to lean towards sicker patients, this also affects the reported death rate. As a consequence, the current data is biased by such restrictions, both in terms of total number of COVID-19 cases in the US which is underestimated, and also death rate due to COVID-19 which is overestimated (5,6).

In spite of the severity of the pandemic, some businesses were asked to maintain productivity during the “stay at home order period” because they are providing an essential function in the US. Lennar Corporation is such a business (Lennar, NYSE: LEN & LEN.B). Lennar is a premier homebuilder (homebuilding is a designated essential function in the US), ~50,000 homes built yearly, present in 21 States and ≥ 78 markets. Lennar employs about 10,000 associates (employees), a cohort that matches well the race and social US distribution. The age range for the company is 18-83 (median age of 49), an age group that covers well over 75% of the COVID19 patients in the US (3). The safety of the associates, customers, and trade partners is the number 1 priority at Lennar, as illustrated by the response of the company to the COVID19 challenge, and such response informs an important knowledge gap relative to industry and the COVID19 pandemic, and might be of interest while businesses are now in the process of reopening.

## METHODS

At the beginning of March, Lennar started collecting real world data via a survey supported by information generated by the CDC guidelines, to ensure the safety of its associates. All 10,000 associates were asked at the beginning of the day and before leaving for work questions about their health specific for COVID-19 (Table 1), based on ethic guidelines provided by the Equal Employment Opportunity Commission (EEOC). This survey, which was conducted using the company’s enterprise resource planning (ERP) system (Workday^®^), started on March 19, 2020, and continued till present, and for a total of 63 consecutive days, or 9 weeks, of data collection. This database was further refined for people whose response was not R1, R2 or R6 (see Table 1), using Lennar’s information extracted from its ERP system (Figure 1). Since the total number of employees is known for the company, the percentage of individuals in each risk category could be calculated. Efforts were made (phone or other contact means) to ensure that associates with no response and those not contacted were not particularly sick or incapacitated, and they were not. Risk categories were developed to rank the severity and associated risk of infection level of the quarantined associates (see Table, individuals who had responded R3, R4 or R5 were asked to answer additional questions about contacts and symptoms via ERP system and phone calls from regional human resource associates).

**Table 1:** Morning Health Check and Stay at Home Information. **<u>Table Legend:</u>** Daily Health Check The Lennar daily survey included eight categorical answers. R0 corresponds to individuals who were contacted but who, for whatever reason, did not respond, we made sure with due followup, that these individuals were not particularly sick or incapacitated, and they were not. R7 corresponds to individuals who, in spite of all efforts, could not be reached. We also ensured that they were not particularly sick or incapacitated. *Direct contact with a COVID-19 patient means physical contact or within 6 feet standing or sitting next at a table/desk; **Indirect contact with a COVID-19 patient means had direct contact with someone who had direct contact with COVID-19 patient, or with a surface presumed to be contaminated by a COVID-19 patient; ***symptoms mean any upper respiratory tract infection symptoms, those typical for COVID-19 (cough, shortness of breath, fever ≥100.4, chills, soar throat, loss of smell/taste, muscle pain), or atypical for COVID-19 (sneezing, runny nose, conjunctivitis, etc.), as further defined by contacting each individual who reported symptoms.

| Categorical Responses on the Digital Daily Morning Health Check at Lennar |
| --- |
| R0: No Response received |
| R1: No symptoms, no contacts or travels in past 14 days, working from home |
| R2: No symptoms, no contacts or travels in past 14 days, working at a work site |
| R3: No symptoms, yes direct contact* or travels in past 14 days, working from home |
| R4: No symptoms, yes indirect contact**, working from home |
| R5: Exhibiting one or more symptoms***, working from home |
| R6: Not scheduled to work today |
| R7: Not contacted |

**Figure 1:**
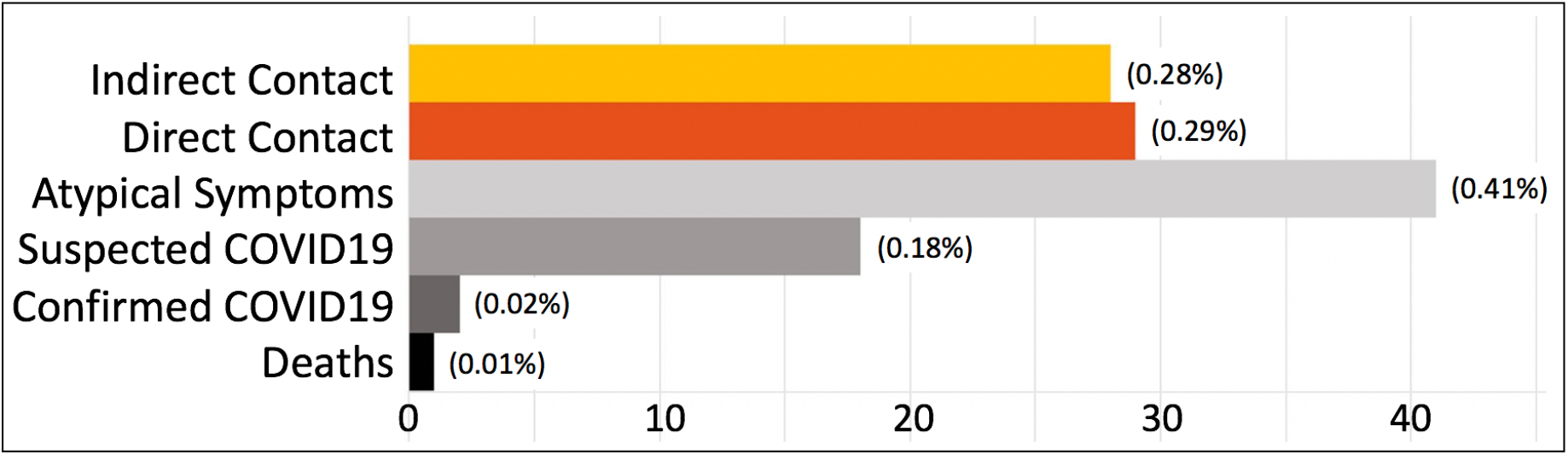
Bar Graph of Counts (and % of total) of Quarantined Associates in Each Risk Category on May 21,2020. Quarantined Associates Per Risk Category (on 5.21.2020) Bar graph showing absolute counts (percentages) per risk category: There were 28 indirect contact; 29 direct contact; 41 atypical airway symptoms for COVID19 (runny nose, sneezing, conjunctivitis, etc.); 18 Suspected COVID-19 patients (CDC Person Under Investigation, not yet tested or awaiting test results) with typical symptoms (see table legend for typical symptoms); There was a total of 14 associates who have tested positive for COVID19 during these 9 weeks, or an average of 1.56 per week. Twelve (12) of these individuals had recovered and were released from quarantine, the others 2 (shown here) were completing their quarantine. One associate was amongst the first six individuals who died of COVID19 in the US, which happened before this database was established, the only associate of Lennar who passed of COVID19.

In addition to this giant survey, many technologies were recruited to ensure the safety, health and wellness of the associates. All associate with symptoms were encouraged to ask their care provider to be tested for COVID19 (testing for COVID19 was done locally, where the associates live, and 100% of the testing was done using the RT-PCR RNA test for SARS-CoV-2). Our real-world experience is that the majority of individuals who have presented with typical COVID-19 symptoms could not be tested until recently (last 3-4 weeks), because of limited test kit supply in the US. All associates with symptoms of upper-airway communicable illnesses were quarantined, 14 days for those “suspected” of COVID19 (one or more typical COVID19 symptoms according to the CDC, and diagnosed as CDC persons under investigation or PUI, by their personal care provider), or “confirmed” with a positive COVID19 test, or 7 days for those suspected of having other upper-airway communicable disorders, based on their atypical symptoms (runny nose, sneezing, conjunctivitis, etc.). Contact tracking was conducted by the completion of a thorough list for anyone known to be infected (positive COVID19 test) or suspected to be infected (CDC PUI) (review of associates, customers, trade partners, family, friends, children, they might have been in contact with). The contact tracking list was then used to request self-quarantine for any associate with direct (14 days) or indirect (7 days) contact (see table legend for definition of direct and indirect contacts) with a COVID19 suspected or test-confirmed individual.

Physical distancing was another intervention, with nearly 50% of the associates working from home (before COVID19, working from home was only available to a small fraction of the associates), and also at work facility using physical distancing (6 feet minimum between associates). The availability of personal protection equipment was implemented, with wearing of a face mask or face cloth, and latex gloves as needed. Hand washing or sanitizing was made available at any work place including in the field. Thorough cleaning of all facilities and new homes with US Environmental Protection Agency-certified products able to destroy CoV-2 were performed regularly, as well as daily scrubbing of surfaces also with EPA-approved COVID19 disinfectants. Additional measures included discontinuation of all in person meetings; quarantine of 14 days for all associates after international travels or cruising, and of four days once back home from domestic travels (or more if they were to develop symptoms); massive information campaign to transparently inform associates of the latest news on the pandemic and the company response (via multimedia: company Video, weekly publications, direct emails for key messages, posters at all work sites, human resource calls, Microsoft, Zoom and other meeting systems, consults on complex issues, etc.), to name just a few measures.

## RESULTS

Based on this cohort of ~10,000, which we continue to survey daily, Lennar was able to generate a database populated by >5,000,000 individual data points over 63 days from March 19 to May 21, 2020 (March 24, 2020 is missing due to IT glitch). The response rate to the daily survey was 88.2% for the first, and 90.8% for the last, five business days of the survey of this period. This real-world database dashboard (Figure 1) showed that 2 associates were quarantined for test-confirmed COVID19 on May 21, 2020. Twelve (12) additional test-confirmed associates, total of 14 during the 63-day survey, had already been returned to work after 14 days of quarantine (or longer), because they were retested for COVID19 and were found to be negative after 14 days (per nasopharyngeal swab and RT-PCR RNA testing for CoV-2), and/or because they were symptom-free for more than 72 hours without symptom-suppressing medications at the end of the quarantine.

On May 21 2020, 18 associates were quarantined at home because they were suspected of having COVID19 (CDC PUI), either because they had typical symptoms, or because their personal care providers had diagnosed them with COVID19. Some of these individuals had been tested (pending results), or not, depending on the availability of COVID19 tests in their geographical area. In addition, 41 associates had been quarantined because of symptoms of other upper respiratory tract infections, symptoms not typically associated with COVID19, and including sneezing/rhinorrhea, conjunctivitis, etc. This group peaked on April 6, and then progressively decreased (Figure 2), and there was very limited opportunity to test this group for COVID19 during the first six weeks of the survey, due to test-kit severe limitation, in spite of our encouragement to get tested.

**Figure 2:**
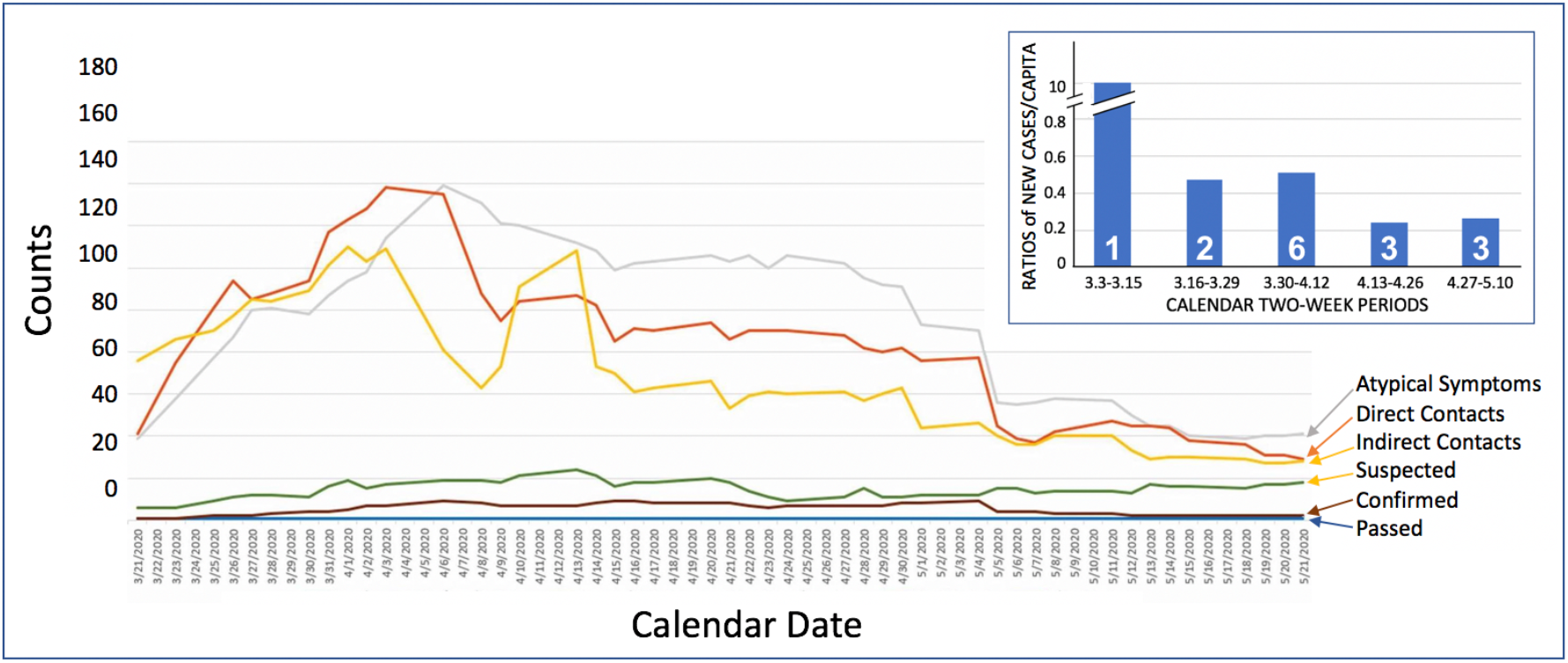
Trend Lines for Quarantined Associates Counts Over Time* and per Risk Category. Trends for Quarantined Associates Line graph recounting the key categories of risk groups, and how they evolved over time. *The *trends* database was started of March 21, not 19. Note that most risk categories peaked between March 30 and April 15, and then decreased thereafter. The total number of COVID19 test-confirmed associates was 14 during that nine-week period, and 12 of the 14 were returned to work after completing their quarantine and were asymptomatic for ≥72hours, and/or were retested by their physicians for COVID19 (nasopharyngeal swab and RT-PCR RNA CoV-2 test) and found to be negative. <u>Inset:</u> Ratios of new cases per capita, Lennar versus total US, showing a progressive reduction of new cases at Lennar compared to the whole US over 5 two-week periods of time, from 3.3 to 5.10, 2020. Number of total COVID19 test-confirmed cases for Lennar corresponding to each two-week period are shown in white at the bottom of each column.

Looking at trends (Figure 2), most of the data have stabilized in April, or even improved, including direct and indirect contacts. These are important figures, direct contact peaked on April 3 and indirect contact peaked on April 1, suggesting that either there were less COVID19 individuals to have contact with, or that associates are less aware of their contacts, or yet other explanations. The positivity-rate of the COVId19 testing has also dropped, from a near 30% at the end of March, to less than 10% in May, 2020. Such recent change is likely explained in part by the increased availability of testing for COVID19 (8,9), but also by less new cases within the company during the second half (4) relative to the first half of the survey (10). Indeed, when new COVID19 cases are expressed as ratios between the count of new cases for Lennar divided by total Lennar associates (10,000), and US count of new cases divided by total US population (330,000,000), for each of five two-week periods, this ratio decreased rather markedly over time (Figure 2, inset). The improvement of Lennar COVID19 progression relative to that of the US may have resulted, at least in part, from our efforts in preventing new infections of our associates with SARS-CoV-2.

## DISCUSSION

### Lessons Learned, and How To Reopen The US Industry Safely

SARS-CoV-2 had two dreadful impacts, first, the deadly consequence on the health of Americans, and second, an unprecedented economic downturn. State governors are struggling with how to prioritize these two scourges, and some governors are reopening businesses in their respective states, because they estimate that the dreadful impact of COVID19 on the “pocketbook of the people” is more significant at this point of the pandemic than the impact on public health (10). How to re-open businesses safely is one challenge that could be helped by our real-world experience at Lennar.

Our database became invaluable in helping the company make essential recommendations. The most important of all was to make sure, using our morning digital health-check, that people who were sick, or simply not “in their usual state of health”, would never come to work and self-quarantine. It turns out, retrospectively, that even if they were to come to work at all and for a few minutes, enough time to just be in a queue and get a temperature check for example, (temperature should be checked at home before going to work), it would be enough contact to markedly increase the risk of contamination of others. The process of self-quarantining for any associate that would represent a contagious risk for others, by making sure that they would not come to any work facility, was <u>our most effective intervention</u>. Associates with symptoms were instructed to isolate themselves at their home and including from their household family members, to minimize viral transmission within their colleagues and the broader communities that they serve (customers, trade partners, communities). It has been shown that even asymptomatic COVID19-infected individuals can contaminate others (11), and when tracking contacts for COVID19 patients, we also searched 48 hours prior to them becoming symptomatic. However, according to our data, we were able to eliminate most contamination of associates at the workplace, due to contact tracking and immediate quarantine of symptomatic COVID19 patients and their (mostly) asymptomatic contacts.

Without feedback from real-world data, it is difficult for any organization to find out if guidelines are being followed and successful in preventing COVID19 infections. Hence, getting data is critical to effective population health management throughout the COVID19 pandemic, as is the ability to test on a large scale the population that we aim to protect, an opportunity that, until recently, has been challenged by limitations in available test kits. We have observed that less than 2% of the associates of our company could be tested, in spite of our effort to encourage our associates to ask their providers for testing, a figure that is, as expected, close to that of the entire US. Data gathering like the morning health checks and testing for COVID19 are both critical to protecting re-opening companies and their employees, and making sure that the protective measures that are now recommended are actually working in protecting people at work. These measures will continue to protect our workforce and communities and hopefully at some point, will be helped by the availability of a successful drug and/or vaccine.

### Conclusion

The data demonstrates that, with appropriate guidelines, and with detailed data, companies like Lennar can continue to be productive, and companies that want to return to productivity, from large corporations to small businesses, could indeed control the proliferation of COVID cases. Strategies pursued by Lennar, if generalized, could minimize viral transmission, and maximize safety, health and wellness within their workforce and the communities they serve.

## Data Availability

All anonymized data is available upon request

## ACKNOWLEDGEMENT

The authors want to thank the leadership team and all the associates of Lennar, whose dedication and commitment to safety, wellness and health made this report possible.TABLE

